# Evaluating post-vaccine expansion patterns of pneumococcal serotypes

**DOI:** 10.1101/2020.05.31.20116228

**Authors:** Maile T. Phillips, Joshua L. Warren, Noga Givon-Lavi, Adrienn Tothpal, Gili Regev-Yochay, Ron Dagan, Daniel M. Weinberger

**Affiliations:** Department of Epidemiology of Microbial Diseases, Yale School of Public Health, New Haven, Connecticut; Department of Biostatistics, Yale School of Public Health, New Haven, Connecticut; Faculty of Health Sciences, Ben-Gurion University of the Negev, Be'er Sheva, Israel; Semmelweird University, Institute of Medical Microbiology, Budapest, Hungary; Sackler Faculty of Medicine, Tel Aviv University, Tel Aviv, Israel; Infection Prevention & Control Unit, Sheba Medical Center, Tel-Hashomer, Israel

**Author notes:** **Corresponding author** Maile T. Phillips, P.O. Box 208034, 60 College St., New Haven, CT 06520-8034 USA.

**Keywords:** pneumococcal conjugate vaccine, pneumococcus, post-vaccine dynamics, *Streptococcus pneumoniae*

## Abstract

*Streptococcus pneumoniae* remains a leading cause of morbidity and mortality. Pneumococcal conjugate vaccines (PCVs) are effective but target only a fraction of the more than 90 pneumococcal serotypes. As a result, the introduction of PCVs has been followed by the emergence of non-vaccine serotypes. With higher-valency PCVs currently under development, there is a need to understand and predict patterns of serotype replacement to anticipate future changes. In this study, we evaluated patterns of change in serotype prevalence post-PCV introduction in Israel. We found that the assumption that non-vaccine serotypes increase by the same proportion overestimates changes in serotype prevalence in Jewish and Bedouin children. Furthermore, pre-vaccine prevalence was positively associated with increases in prevalence over the study period. From our analyses, serotypes 12F, 8, 16F, 33F, 9N, 7B, 10A, 22F, 24F, and 17F were estimated to have gained the most cases of invasive pneumococcal disease through serotype replacement in the Jewish population. However, this model also failed to quantify some additional cases gained, suggesting that changes in carriage in children alone may be insufficient to explain serotype replacement in disease. Understanding of serotype replacement is important as higher-valency vaccines are introduced.

---

*Streptococcus pneumoniae* (pneumococcus) is a leading cause of morbidity and mortality globally and causes a range of diseases including pneumonia, meningitis, and septicemia [1]. Nasopharyngeal carriage of pneumococcus is a precursor for disease [2]. Carriage is common in young children, who are the main reservoir of transmission to adults and other children [2, 3]. The likelihood of developing disease following exposure to pneumococcus depends on bacterial factors (i.e., serotype and other virulence factors) and host factors (age, immunocompetence, recent viral infection) [4, 5].

Pneumococcal conjugate vaccines (PCVs) protect against carriage and invasive infections due to serotypes targeted by the vaccine [6–9]. However, PCVs target only a fraction of the more than 90 pneumococcal serotypes. As a result, despite an overall decline in pneumococcal disease, the introduction of PCVs is followed by the emergence of non-vaccine serotypes among healthy carriers and, consequently, as causes of disease [6, 10, 11]. This ‘serotype replacement’ has reduced the overall impact of PCVs on disease rates and has prompted the development of PCVs that target larger numbers of serotypes. Several new PCVs are under development that target 15, 20, or more serotypes [12–14]. These vaccines will again likely disrupt the balance of serotypes in the nasopharynx and lead to further serotype replacement. As these next-generation conjugate vaccines move towards licensure, it is important to understand likely patterns of serotype replacement so that the marginal benefit of higher-valency vaccines can be anticipated. This is useful as countries decide which product to recommend in a given setting.

When evaluating the potential impact of new vaccines, there is a need to predict the overall benefits, which are influenced by both declines in disease incidence due to vaccine-targeted serotypes and by increases in disease incidence due to non-vaccine serotypes. At the same time, it is important to predict which serotypes are likely to emerge to help with future adjustments to the vaccine. To produce such projections, it is necessary to make assumptions about which serotypes are likely to increase and by how much. There are several proposed frameworks for accomplishing this goal. Nurhonen and Auranen predicted the impact of alternative PCV formulations with the assumption that all non-vaccine serotypes increase by the same proportion following vaccine introduction and that the invasiveness of the non-vaccine serotypes as a group is constant over time [15]. This approach can successfully capture the overall magnitude of the benefit of a vaccine by lumping vaccine and non-vaccine serotypes into broad groups [16–18]. However, when the goal is to examine emerging serotypes, it is important to consider individual serotypes. Recent work has used genomic data with models of negative frequency-dependent selection to quantify post-vaccine expansion patterns [19, 20]. However, whole genome sequence data are not always widely available, and their interpretation is complex. A complementary approach would be to identify characteristics of serotypes that emerge following vaccine introduction and to use this information to inform projections.

In this study, we characterized the expansion patterns of non-vaccine serotypes among healthy carriers following the introduction of PCVs in Israel. We evaluated correlates of serotype expansion, compared the expansion patterns in Israel to those that would be expected if the commonly-employed assumption that all non-vaccine serotypes expand by the same amount were correct, and assessed the implications for disease projections if this assumption is wrong. Finally, we provided a framework to rank serotypes not currently included in PCV13 based on the likelihood that they may be important causes of disease in the future.

## METHODS

### Data, setting, & participants

PCV7 was introduced for use in Israel in the national immunization program in July 2009 with a catch-up program in children <2 years old and was replaced by PCV13 in November 2010 without further catch-up. Carriage data were collected as part of prospective surveillance in southern Israel from November 2009-June 2016, described in detail elsewhere [21]. Every weekday during the study period, nasopharyngeal cultures were collected from the first four Jewish and first four Bedouin children under five presenting at a pediatric emergency department in Be’er Sheva, Israel for any reason. Information was recorded regarding ethnicity, result of the pneumococcal culture, pneumococcal serotype (if applicable), clinical diagnosis, number of PCV doses received to date (PCV7 and/or PCV13), and the year and month when the swab was recorded.

Invasive pneumococcal disease (IPD) data were also collected during the same time period as part of nationwide surveillance. These data were collected for individuals of all ages. Variables were collected regarding age group (<5, 5–17, 18–39, 40–64, and 65+ years), serotype, date (July 2009- June 2016), and number of swabs. Both the carriage and IPD datasets were aggregated into counts by serotype and epidemiological year (seven July-June years); the number of swabs negative for pneumococcus in the carriage dataset was also tallied. Year 0 in the carriage dataset is only ¾ of an epidemiological year, due to the fact that data collection started in November (instead of July) that year.

### Other data

Additional serotype-specific characteristics were used in regression models to evaluate associations with baseline carriage prevalence and with changes in prevalence. These serotype-specific variables included relative density of growth reached at an early time point during *in vitro* growth [22], case fatality rate, negative charge carbon, total carbon per polysaccharide repeat, and capsule thickness [23]. In instances of missing data, missing values were imputed in the model assuming the data were missing completely at random (S1 Text).

### Carriage model

The goal was to quantify changes in the prevalence of carriage of individual serotypes and to evaluate correlates of baseline prevalence or changes in prevalence. To do this, we fit a hierarchical Bayesian regression model to the data. The data for individual serotypes were sparse, so we used a hierarchical prior structure to provide stabilized estimates of the prevalence ratios for all serotypes [24]. In this model, pneumococcal carriage at time (epidemiological year) *t* follows a multinomial distribution with the number of successes being the number of detections of each non-vaccine serotype (i.e., each non-vaccine serotype representing a unique category), and the number of trials being the total number of nasopharyngeal swabs at time *t*. The “reference” category in this model includes all negative swabs and detections of PCV13 serotypes combined. The change in prevalence in the post-vaccine period varies by serotype and is a function of serotype-specific covariates. In this model, time is defined as a three-category variable, with the three categories corresponding to the pre- (November 2009-June 2010), early post-PCV (July 2010-June 2012), and late post-PCV (July 2012-June 2016) vaccination periods. This time structure provided a better fit compared to linear time, and was compared to several other functions of time. The three-category structure of time allows serotype prevalence to level out to its final post-vaccine prevalence, facilitating a better comparison of pre- to post-vaccine prevalence. The overall model is defined as:

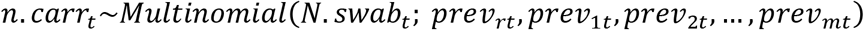

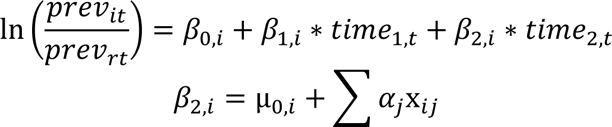

*n.carr_t_* = pneumococcal carriage count at time period *t*
*N.swab_t_* = total # swabs at time period *t*
*previ_it_* = pneumococcal prevalence for non-vaccine serotype *i* at time period *t* (m=54 non-vaccine serotypes)
*previ_rt_* =pneumococcal prevalence from the “reference” group at time period *t* (“reference” swab is defined as negative for any pneumococcus or positive for vaccine-serotypes)
*i* = 1, 2,…, m
*t*= 0, 1,…, 6 (year)
*time1*= 0, 1, 1, 1, 1, 1, 1
*time2*= 0, 0, 0, 1, 1, 1, 1
x*_ij_*= the *j*_th_ serotype-level covariate for non-vaccine serotype *i*

All prior distributions were chosen to be weakly informative. The intercepts (relative prevalence at *time 0*) and slopes (early and late post-vaccine prevalence compared to *time 0*) were allowed to vary by serotype and were estimated hierarchically by having the serotype-specific parameters centered around global parameters that were common to all non-vaccine serotypes. Conjugate priors were used in all cases except for the standard deviation parameters, where uniform priors were used [25].

Different assumptions about model structure and covariates were evaluated for this model. Variations in random and fixed effects were considered, different structures of time were evaluated, and covariance matrices were applied to model both independence and dependence of the intercepts and slopes. The best model formulation was chosen using the optimal deviance information criterion (DIC) [26], using decreases of at least 10 to be considered an improved model. Using Markov chain Monte Carlo sampling techniques, we obtained 150,000 posterior samples from the joint posterior distribution following a burn-in period of 10,000 iterations. The Gelman-Rubin [27] and Geweke [28] diagnostics were used to investigate convergence of individual parameters and effective sample sizes were calculated to ensure we collected enough posterior samples post-convergence to make accurate inference. Additional details about the model, including structure and model diagnostics, can be found in the supplementary text (S1 Text).

Serotype-specific prevalence, relative risk ratios (RRRs; the multiplicative change for a one-year increase for a specific serotype prevalence relative to the reference prevalence), slopes, and intercepts were estimated from the final model. Where relevant, these estimates were compared to the corresponding values from the constant proportional change model, which assumes that serotype replacement in carriage is complete and that all serotypes increase by the same factor (i.e., prevalence ratio is the same for all serotypes).

### Linking carriage with disease

We sought to quantify the impact of serotype replacement on IPD. This is challenging because many serotypes have secular trends or exhibit epidemic patterns even in the absence of the vaccines. Simply comparing the cases of IPD before and after vaccination can sometimes be misleading. Instead, we chose a more stable approach which used the estimates of the serotype-specific prevalence ratios from our carriage model to estimate the additional cases gained as a result of increased carriage in children (*cases.gained.explained*). The number of cases that would be expected in the absence of changes in carriage in children is calculated by dividing the observed number of cases at a given time point by the prevalence ratio for the same serotype. The cases gained (explained by increased carriage) estimate is calculated by subtracting this counterfactual for the number of cases from the observed number of cases for each serotype, age category and time point:

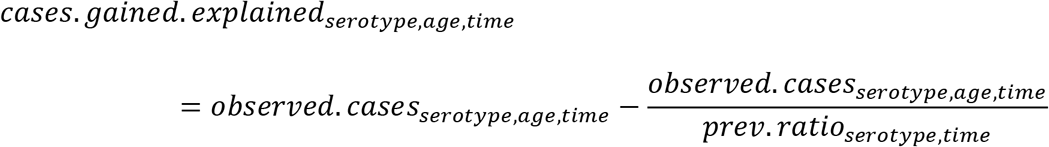

where the prevalence ratios (*prev.ratio*) from *time 0* to *time 6* were estimated from the model. We also calculated the number of cases gained that were not explained by increased carriage in children (e.g., could be due to secular trends; *cases.gained.unexplained*), by subtracting the observed number of IPD cases at *time 0* from the number of cases that would have been expected in the absence of changes in carriage in children (i.e., the number of IPD cases in the first time period):

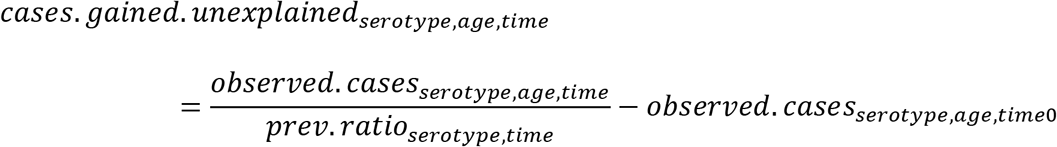

Using the values of cases gained as a result of increased carriage in children, we sorted the serotypes from highest to lowest. All of these calculations were carried out using results from the Jewish carriage model only, since the IPD dataset predominantly included Jewish individuals.

In some instances, the expected number of IPD cases exceeded the number of observed cases. In such cases, we adjusted the expected number of cases to be the number of observed cases. In other words, we let the maximum number of expected cases be the number of observed cases, resulting in zero cases gained in that instance.

To assess whether changes in carriage in younger children or older children better correlated with changes in IPD in adults, we additionally fit the carriage model separately for younger children (<12 months) and older children (different subsets of 24–60-month-old children), and compared the number of unexplained vs explained cases gained between the two age groups.

All analyses were performed using R version 3.4.0 [29]. The carriage model was fit using JAGS version 4.3.0 [30]. All code, with simulated data, can be found online at https://github.com/mailephillips/post-pcv-expansion.

## RESULTS

### Characteristics of the data

The Israel carriage dataset contained 10,396 observations, with 57.1% of samples from Bedouin children and the rest from Jewish children. Of all swabs, 47.7% were positive for pneumococcus, with 42.6% positive among Jewish children and 51.5% positive among Bedouin children. In 2009, just after vaccine introduction, 63.5% of colonized children had a vaccine-targeted serotype and 36.5% had a non-vaccine serotype. The most commonly-carried non-PCV13 serotypes in 2009 were 15B/C, 15A, 16F, 10A, 38, 35B, 23B, 23A, 21, and 10B.

### Serotype expansion patterns in pneumococcal carriage

Some typical assumptions when projecting serotype replacement are that all non-vaccine serotypes increase by the same amount (i.e., same prevalence ratio), that the rank order of non-vaccine serotypes is maintained following vaccine introduction, and that non-vaccine serotypes completely replace vaccine serotypes in carriage. When we compare the observed pre-vaccine prevalence of serotypes to the post-vaccine prevalence, serotypes increase by different proportions and do not maintain rank order, suggesting that the assumption of constant prevalence ratio does not hold (Figure 1). However, our analysis suggests that instead some serotypes expand more than expected, and others remain stable or even decline (Figure 2).

**Figure 1.**
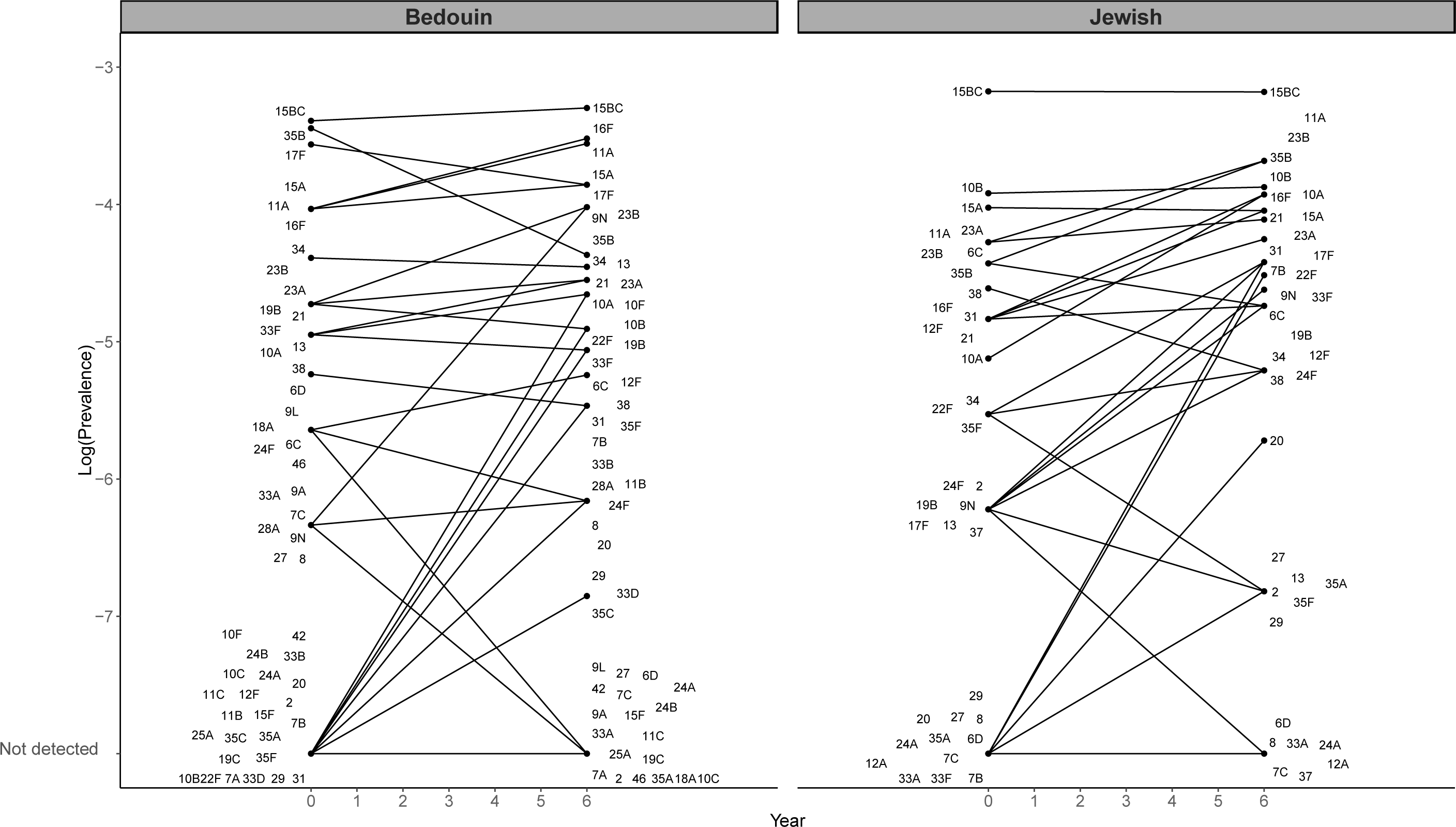
Pre-vaccine vs. post-vaccine pneumococcal prevalence of non-vaccine types for children under 5 in Israel, November 2009-July 2016. The observed log-prevalence of pneumococcus for each serotype is shown at Year 0 and Year 6, separately for Jewish and Bedouin children. If a serotype increased from or decreased to 0, the log prevalence is shown as “not detected” at the bottom of the graph.

**Figure 2.**
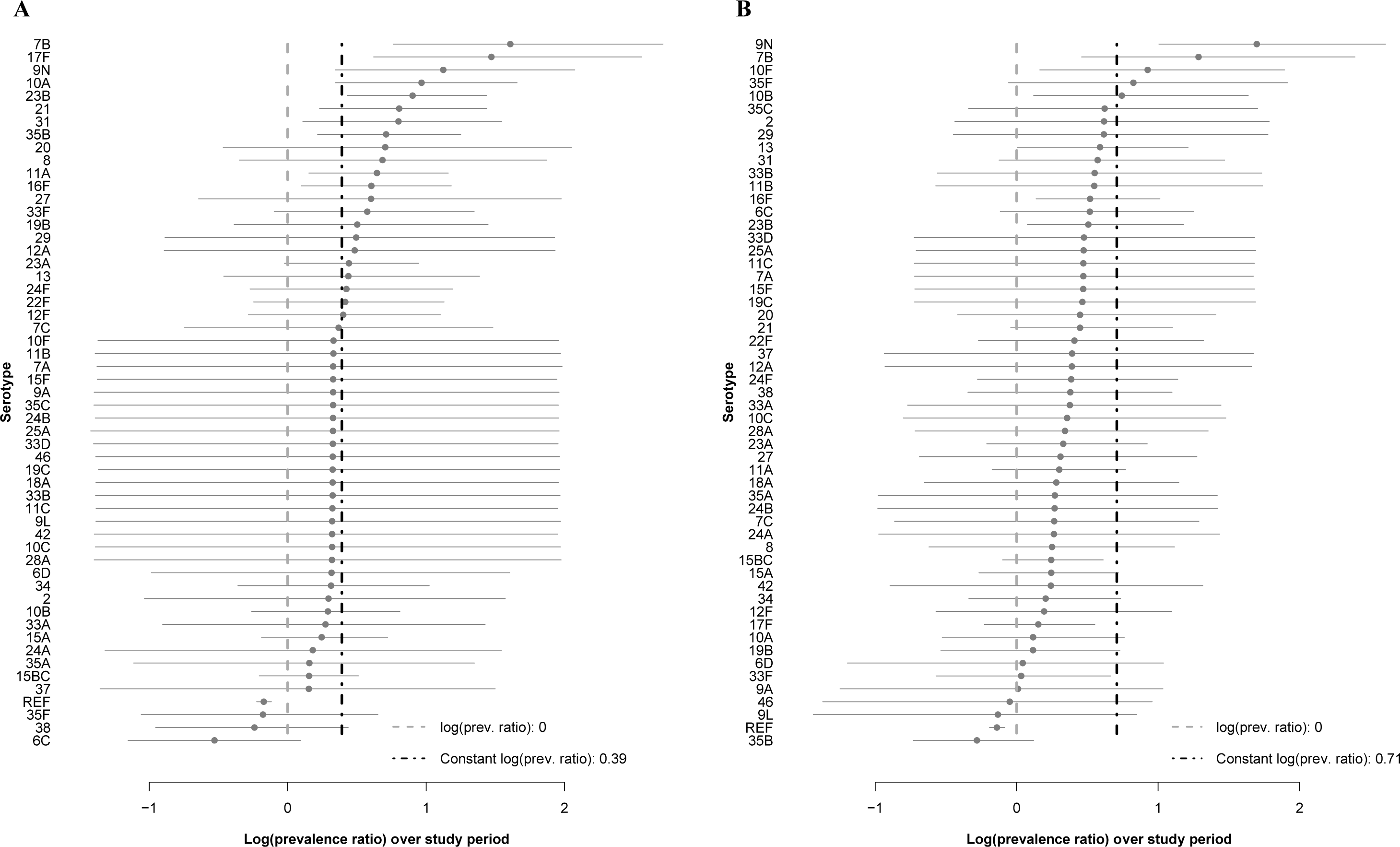
Log(prevalence ratios) from the carriage models for children under 5 in Israel, November 2009-June 2016. Log-transformed ratios comparing late post- to pre-vaccine prevalence (last year/first year) are shown for each serotype with 95% credible intervals from the Jewish and Bedouin carriage models. The serotypes are shown from highest to lowest log prevalence ratios.

The ten serotypes with the largest relative increase in carriage prevalence among Jewish children, from highest to lowest, were 7B, 17F, 9N, 10A, 23B, 21, 31, 35B, 20, and 8. Among Bedouin children, the largest relative increases were 9N, 7B, 10F, 35F, 10B, 35C, 2, 29, 13, and 31 (Figure 2, Table S2). The majority of the prevalence ratios were larger than one in both models (Jewish median prevalence ratio: 1.35, Bedouin median: 1.36). In both models, many serotypes had changes that were smaller than expected based on the model assuming proportional increases (constant proportional increase: 1.48 Jewish, 2.03 Bedouin). The serotype-specific intercepts and other serotype-specific parameters estimated from the model can be found in Table S2 and Figures S5–7.

### Correlates of serotype-specific changes

While serotype-specific prevalence of the non-PCV13 serotypes mostly increased after vaccination, the degree to which each serotype changed varied. The distribution of the change in prevalence post-vaccination was skewed, but we were able to predict where on the distribution each serotype was located using serotype-specific characteristics. We evaluated several serotype-specific correlates of fitness to try to explain this pattern. The best correlate of the post-vaccine increase was pre-vaccine prevalence. The relationship between pre-vaccine prevalence and the change in serotype prevalence over time improved model fit when included with a covariance matrix compared to all other combinations of predictors. When fit to carriage data from Jewish children, the model with the covariance matrix was the best model, based on DIC. When fit to carriage data from Bedouin children, the model with independent slopes (no predictors) and the model with the covariance matrix both fit the data best based on DIC. There was an overall positive association between pre-vaccine and the change to late post-PCV prevalence in the Jewish dataset (0.04, 95% credible interval: –0.68, 0.68), and a negative association in the Bedouin dataset (−0.21, 95% credible interval: –0.69, 0.34). Combinations of relative density, case fatality rate, negative charge carbon, total carbon per polysaccharide repeat, and capsule thickness were not as strongly associated with the prevalence ratio and resulted in worse model fits in both datasets.

### Ranking serotypes based on estimated cases gained

From 2009–2015, there were 3,968 total cases of IPD. We estimate that during this period, there was an excess of 624 (95% credible interval: 397, 850) cases gained across all age groups as a result of increased carriage of non-vaccine serotypes in children (Figure 3, Table 1). From highest to lowest, serotypes 12F, 8, 16F, 33F, 9N, 7B, 10A, 22F, 24F, and 17F were estimated to have gained the most cases of IPD through serotype replacement (each with an estimated 21 to 112 additional cases gained). These 10 serotypes combined had a total of 1,223 reported IPD cases during this period. An estimated 446 of these cases could be attributed to increases in carriage prevalence (Table 1). Fourteen serotypes did not account for any additional cases gained across this study period.

**Figure 3.**
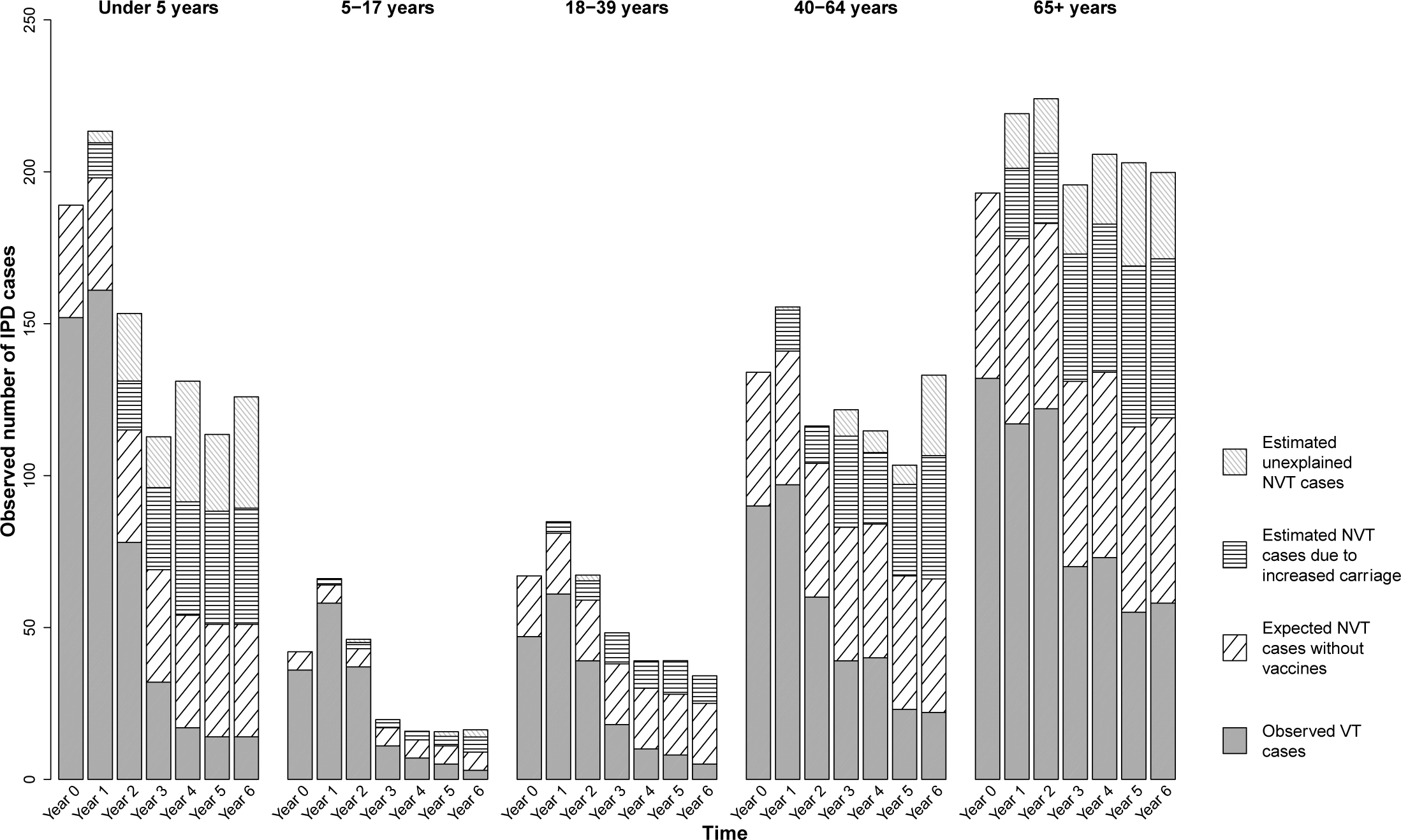
Estimated additional cases gained as a result of increased carriage in children under 5 in Israel, July 2009-July 2016. Results are shown for each age group and year of the study. Each full bar represents the total number of observed IPD cases for that stratum. The different patterned portions of the bars denote the fraction of total cases attributable to different groups of serotypes: observed vaccine-targeted (VT) serotype cases (solid gray), expected non-vaccine-targeted (NVT) serotype cases had PCV13 not been introduced (diagonal black lines), estimated additional cases gained as a result of increased carriage in children, explained by this model (horizontal black lines), and estimated additional cases gained that are not explained by this model (gray diagonal gray lines).

**Table 1:** Model Estimates and Cases Gained Through Serotype Replacement over 6 Years among Jewish Individuals in Israel (July 2009-June 2016), Ranked from Most to Least Cases Gained in All Age Categories. The results of the observed carriage at time 0, observed prevalence ratio (time6/time0), model prevalence ratio (time6/time0), and estimated cases gained through serotype replacement by age groups are shown for each of the serotypes used in the carriage model. Serotypes are ordered from most to least cases gained among all ages. Note that the total cases gained in the last row adds up to more than the number of cases above for each serotype, because the serotypes shown are only those used in the carriage model. The values for total cases gained also included calculations for additional serotypes (not shown) that were responsible for IPD, but were not present in carriage in children under 5 during this time period.

| Serotype | Observed |  | Model |  | Cases gained over study period, by age in years |  |  |  |  |  |  |  |  |  |  |  |
| --- | --- | --- | --- | --- | --- | --- | --- | --- | --- | --- | --- | --- | --- | --- | --- | --- |
|  | <5<br>carr.<br>(t<sub>0</sub>) | PR<br>(t<sub>6</sub>/t<sub>0</sub>) | PR (t<sub>6</sub>/t<sub>0</sub>) |  | <5 |  | 5-17 |  | 18-39 |  | 40-64 |  | 65+ |  | All Ages |  |
|  |  |  | Med. | CrI | Med. | CrI | Med. | CrI | Med. | CrI | Med. | CrI | Med. | CrI | Med. | CrI |
| 12F | 4 | 1.10 | 1.48 | 0.75, 2.98 | 58 | 0, 114 | 3 | 0, 6 | 8 | 0, 15 | 20 | 0, 39 | 22 | 0, 44 | 112 | 0, 219 |
| 8 | 0 | NA** | 1.96 | 0.69, 6.44 | 4 | 0, 7 | 3 | 0, 5 | 14 | 0, 23 | 29 | 0, 49 | 23 | 0, 38 | 72 | 2, 122 |
| 16F | 4 | 2.48 | 1.82 | 1.10, 3.29 | 6 | 1, 9 | 0 | 0, 0 | 3 | 1, 4 | 14 | 4, 24 | 27 | 7, 45 | 50 | 13, 82 |
| 33F | 0 | NA* | 1.78 | 0.90, 3.95 | 20 | 1, 35 | 0 | 0, 1 | 3 | 0, 5 | 5 | 1, 9 | 11 | 1, 19 | 39 | 4, 69 |
| 9N | 1 | 1.00 | 3.08 | 1.39, 8.09 | 5 | 2, 7 | 1 | 1, 2 | 4 | 2, 5 | 9 | 4, 12 | 15 | 6, 20 | 35 | 15, 46 |
| 7B | 0 | NA* | 4.98 | 2.13, 15.25 | 10 | 6, 11 | 0 | 0, 0 | 1 | 1, 1 | 6 | 4, 7 | 15 | 10, 18 | 32 | 21, 38 |
| 10A | 3 | 3.30 | 2.62 | 1.40, 5.28 | 10 | 4, 14 | 0 | 0, 0 | 2 | 1, 2 | 8 | 4, 11 | 11 | 5, 15 | 31 | 14, 42 |
| 22F | 2 | 3.03 | 1.50 | 0.79, 3.10 | 5 | 0, 11 | 1 | 0, 2 | 2 | 0, 3 | 9 | 0, 17 | 11 | 0, 22 | 28 | 1, 55 |
| 24F | 1 | 2.75 | 1.52 | 0.75, 3.32 | 7 | 0, 15 | 1 | 0, 2 | 2 | 0, 4 | 6 | 0, 12 | 12 | 0, 24 | 28 | 1, 56 |
| 17F | 1 | 6.05 | 4.32 | 1.84, 13.00 | 3 | 2, 4 | 0 | 0, 1 | 2 | 1, 3 | 5 | 3, 6 | 10 | 6, 13 | 21 | 12, 27 |
| 23B | 6 | 2.11 | 2.46 | 1.52, 4.19 | 4 | 2, 5 | 2 | 1, 2 | 0 | 0, 1 | 4 | 2, 5 | 12 | 7, 15 | 21 | 12, 28 |
| 15A | 9 | 0.98 | 1.28 | 0.82, 2.06 | 3 | 0, 8 | 0 | 0, 1 | 0 | 0, 0 | 4 | 0, 9 | 13 | 0, 28 | 21 | 0, 46 |
| 31 | 4 | 1.79 | 2.21 | 1.10, 4.62 | 1 | 0, 1 | 1 | 0, 1 | 1 | 0, 2 | 6 | 1, 8 | 12 | 3, 19 | 21 | 4, 31 |
| 11A | 7 | 1.81 | 1.90 | 1.17, 3.22 | 2 | 0, 3 | 1 | 0, 1 | 2 | 1, 4 | 7 | 2, 10 | 8 | 2, 12 | 19 | 6, 29 |
| 35B | 6 | 2.11 | 2.03 | 1.23, 3.50 | 2 | 1, 3 | 0 | 0, 0 | 1 | 0, 1 | 3 | 1, 5 | 11 | 4, 16 | 17 | 6, 25 |
| 15BC | 21 | 1.00 | 1.16 | 0.81, 1.66 | 8 | 0, 20 | 0 | 0, 0 | 1 | 0, 2 | 3 | 0, 7 | 4 | 0, 10 | 16 | 0, 39 |
| 10B | 10 | 1.05 | 1.33 | 0.76, 2.27 | 5 | 0, 10 | 0 | 0, 1 | 0 | 0, 1 | 1 | 0, 2 | 6 | 0, 13 | 12 | 0, 27 |
| 2 | 1 | 0.55 | 1.32 | 0.35, 4.71 | 3 | 0, 8 | 2 | 0, 6 | 2 | 0, 6 | 3 | 0, 11 | 1 | 0, 3 | 11 | 0, 34 |
| 23A | 7 | 1.18 | 1.55 | 0.97, 2.61 | 1 | 0, 2 | 0 | 0, 0 | 0 | 0, 1 | 4 | 0, 7 | 5 | 0, 8 | 10 | 1, 17 |
| 34 | 2 | 1.38 | 1.36 | 0.69, 2.80 | 1 | 0, 1 | 0 | 0, 1 | 0 | 0, 1 | 1 | 0, 2 | 6 | 0, 13 | 7 | 0, 17 |
| 27 | 0 | NA* | 1.83 | 0.52, 7.33 | 6 | 0, 12 | 0 | 0, 1 | 0 | 0, 0 | 0 | 0, 0 | 0 | 0, 0 | 6 | 0, 12 |
| 38 | 5 | 0.55 | 0.78 | 0.38, 1.56 | 1 | 0, 4 | 0 | 0, 0 | 0 | 0, 0 | 1 | 0, 4 | 3 | 0, 10 | 5 | 0, 17 |
| 13 | 1 | 0.55 | 1.54 | 0.62, 4.00 | 0 | 0, 1 | 0 | 0, 1 | 0 | 0, 0 | 2 | 0, 4 | 2 | 0, 5 | 5 | 0, 10 |
| 21 | 4 | 2.20 | 2.22 | 1.25, 4.20 | 3 | 1, 5 | 0 | 0, 0 | 0 | 0, 0 | 0 | 0, 0 | 0 | 0, 0 | 3 | 1, 5 |
| 6C | 6 | 0.73 | 0.58 | 0.32, 1.09 | 1 | 0, 2 | 0 | 0, 0 | 0 | 0, 1 | 0 | 0, 2 | 2 | 0, 5 | 3 | 0, 9 |
| 20 | 0 | NA* | 1.99 | 0.62, 7.66 | 0 | 0, 0 | 0 | 0, 0 | 1 | 0, 1 | 1 | 0, 1 | 2 | 0, 3 | 3 | 0, 5 |
| 29 | 0 | NA* | 1.60 | 0.40, 6.86 | 1 | 0, 3 | 0 | 0, 0 | 0 | 0, 0 | 0 | 0, 0 | 0 | 0, 1 | 1 | 0, 3 |
| 35F | 2 | 0.28 | 0.83 | 0.34, 1.90 | 0 | 0, 1 | 0 | 0, 0 | 0 | 0, 1 | 0 | 0, 1 | 1 | 0, 5 | 1 | 0, 8 |
| 9A | 0 | NA** | 1.35 | 0.24, 6.93 | 0 | 0, 1 | 0 | 0, 0 | 0 | 0, 1 | 0 | 0, 1 | 0 | 0, 1 | 1 | 0, 3 |
| 24B | 0 | NA** | 1.34 | 0.23, 6.87 | 1 | 0, 2 | 0 | 0, 0 | 0 | 0, 0 | 0 | 0, 1 | 0 | 0, 0 | 1 | 0, 3 |
| 18A | 0 | NA** | 1.35 | 0.23, 6.77 | 0 | 0, 0 | 0 | 0, 0 | 0 | 0, 0 | 1 | 0, 2 | 0 | 0, 1 | 1 | 0, 3 |
| 25A | 0 | NA** | 1.35 | 0.24, 6.75 | 0 | 0, 0 | 0 | 0, 0 | 0 | 0, 0 | 0 | 0, 1 | 0 | 0, 1 | 1 | 0, 2 |
| 10F | 0 | NA** | 1.35 | 0.24, 6.91 | 0 | 0, 0 | 0 | 0, 0 | 0 | 0, 1 | 0 | 0, 0 | 0 | 0, 1 | 1 | 0, 2 |
| 33A | 0 | NA** | 1.29 | 0.40, 4.06 | 0 | 0, 1 | 0 | 0, 0 | 0 | 0, 0 | 0 | 0, 0 | 0 | 0, 0 | 0 | 0, 1 |
| 19B | 1 | 4.40 | 1.63 | 0.67, 4.23 | 0 | 0, 0 | 0 | 0, 0 | 0 | 0, 1 | 0 | 0, 0 | 0 | 0, 0 | 0 | 0, 1 |
| 6D | 0 | NA** | 1.36 | 0.36, 4.87 | 0 | 0, 0 | 0 | 0, 0 | 0 | 0, 0 | 0 | 0, 0 | 0 | 0, 1 | 0 | 0, 1 |
| 9L | 0 | NA** | 1.35 | 0.23, 7.02 | 0 | 0, 0 | 0 | 0, 0 | 0 | 0, 0 | 0 | 0, 1 | 0 | 0, 0 | 0 | 0, 1 |
| 37 | 1 | 0.00 | 1.13 | 0.25, 4.35 | 0 | 0, 0 | 0 | 0, 0 | 0 | 0, 1 | 0 | 0, 1 | 0 | 0, 0 | 0 | 0, 2 |
| 28A | 0 | NA** | 1.35 | 0.24, 6.83 | 0 | 0, 0 | 0 | 0, 0 | 0 | 0, 0 | 0 | 0, 0 | 0 | 0, 1 | 0 | 0, 1 |
| 11C | 0 | NA** | 1.34 | 0.23, 6.88 | 0 | 0, 0 | 0 | 0, 0 | 0 | 0, 0 | 0 | 0, 0 | 0 | 0, 1 | 0 | 0, 1 |
| 35A | 0 | NA* | 1.16 | 0.32, 3.77 | 0 | 0, 0 | 0 | 0, 0 | 0 | 0, 0 | 0 | 0, 0 | 0 | 0, 1 | 0 | 0, 1 |
| 10C | 0 | NA** | 1.34 | 0.24, 6.90 | 0 | 0, 0 | 0 | 0, 0 | 0 | 0, 0 | 0 | 0, 0 | 0 | 0, 0 | 0 | 0, 0 |
| 11B | 0 | NA** | 1.34 | 0.24, 6.85 | 0 | 0, 0 | 0 | 0, 0 | 0 | 0, 0 | 0 | 0, 0 | 0 | 0, 0 | 0 | 0, 0 |
| 12A | 0 | NA** | 1.60 | 0.40, 6.97 | 0 | 0, 0 | 0 | 0, 0 | 0 | 0, 0 | 0 | 0, 0 | 0 | 0, 0 | 0 | 0, 0 |
| 15F | 0 | NA** | 1.35 | 0.24, 7.00 | 0 | 0, 0 | 0 | 0, 0 | 0 | 0, 0 | 0 | 0, 0 | 0 | 0, 0 | 0 | 0, 0 |
| 19C | 0 | NA** | 1.34 | 0.24, 6.88 | 0 | 0, 0 | 0 | 0, 0 | 0 | 0, 0 | 0 | 0, 0 | 0 | 0, 0 | 0 | 0, 0 |
| 24A | 0 | NA** | 1.17 | 0.26, 4.59 | 0 | 0, 0 | 0 | 0, 0 | 0 | 0, 0 | 0 | 0, 0 | 0 | 0, 0 | 0 | 0, 0 |
| 33B | 0 | NA** | 1.35 | 0.24, 6.87 | 0 | 0, 0 | 0 | 0, 0 | 0 | 0, 0 | 0 | 0, 0 | 0 | 0, 0 | 0 | 0, 0 |
| 33D | 0 | NA** | 1.35 | 0.24, 6.84 | 0 | 0, 0 | 0 | 0, 0 | 0 | 0, 0 | 0 | 0, 0 | 0 | 0, 0 | 0 | 0, 0 |
| 35C | 0 | NA** | 1.34 | 0.24, 6.89 | 0 | 0, 0 | 0 | 0, 0 | 0 | 0, 0 | 0 | 0, 0 | 0 | 0, 0 | 0 | 0, 0 |
| 42 | 0 | NA** | 1.34 | 0.24, 6.86 | 0 | 0, 0 | 0 | 0, 0 | 0 | 0, 0 | 0 | 0, 0 | 0 | 0, 0 | 0 | 0, 0 |
| 46 | 0 | NA** | 1.36 | 0.24, 6.93 | 0 | 0, 0 | 0 | 0, 0 | 0 | 0, 0 | 0 | 0, 0 | 0 | 0, 0 | 0 | 0, 0 |
| 7A | 0 | NA** | 1.34 | 0.23, 6.84 | 0 | 0, 0 | 0 | 0, 0 | 0 | 0, 0 | 0 | 0, 0 | 0 | 0, 0 | 0 | 0, 0 |
| 7C | 0 | NA** | 1.42 | 0.47, 4.39 | 0 | 0, 0 | 0 | 0, 0 | 0 | 0, 0 | 0 | 0, 0 | 0 | 0, 0 | 0 | 0, 0 |
| All |  |  |  |  | 167 | 88, 245 | 17 | 10, 26 | 49 | 29, 70 | 149 | 94, 203 | 242 | 162, 320 | 624 | 397, 850 |

carr.=carriage; CrI=95% credible interval; med.=median; PR=prevalence ratio.
a \* Indicates that the prevalence at Time 0 was 0, and hence the observed prevalence ratio could not be estimated for that serotype. In instances where the prevalence at Time 6 was also 0, there is an additional asterisk (\*\*).

Serotype 12F, in particular, was responsible for 112 cases gained as a result of increased carriage in children out of the total 394 IPD cases for 12F. In all age groups, 12F was among the three highest ranked serotypes among cases gained as a result of increased carriage in children. Serotype 8 was similarly highly ranked for cases gained as a result of increased carriage in children in all age groups except for children <5 years. Serotype 16F was highly ranked among cases gained among adults 18 years and older, driving up its overall ranking.

Adults 65+ and children under five years were responsible for the most IPD cases (Table 1, Figure 3). In general, observed vaccine-type cases decreased over the study period, whereas the additional cases gained (both explained and unexplained) increased over the study period. Plots for individual serotype IPD cases over the study period can be found in Figure S8. Serotype 12F in particular was responsible for the most estimated cased gained that are not explained by increased carriage in children.

We had intended to compare results from carriage models with children less than one year and different subsets of 24–60 months of age to assess whether changes in carriage in younger children or older children better correlated with changes in IPD in adults; however, the data were too sparse for a formal comparison.

### Cases not explained by carriage

The number of additional cases gained over this time period that were not explained by increased carriage in children was also substantial. Over the study period, there were an estimated 462 (95% credible interval: 312, 641) cases that were not predicted based on changes in carriage in children. The serotypes with the most unexplained cases gained were 12F, 24F, 22F, 10A, and 15BC (Table 2). Serotype 12F had the most unexplained cases, with an estimated 156 (95% credible interval: 50, 268) cases.

**Table 2.**
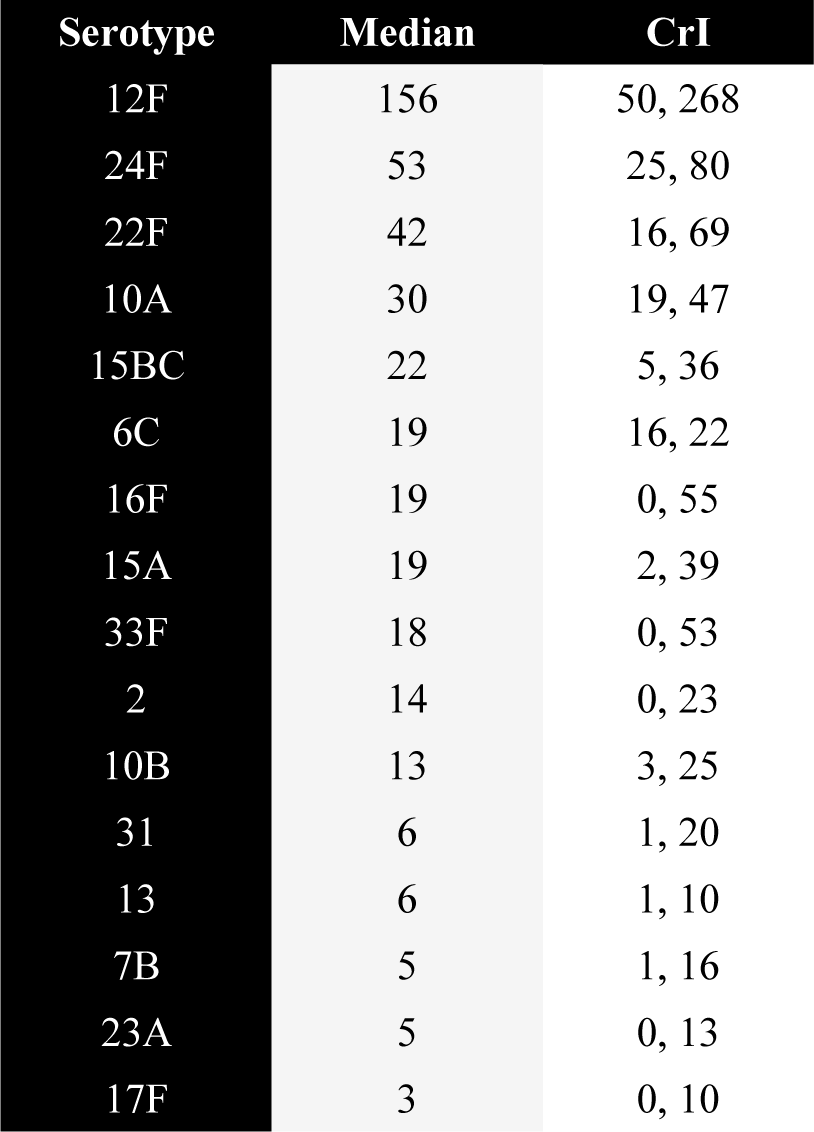
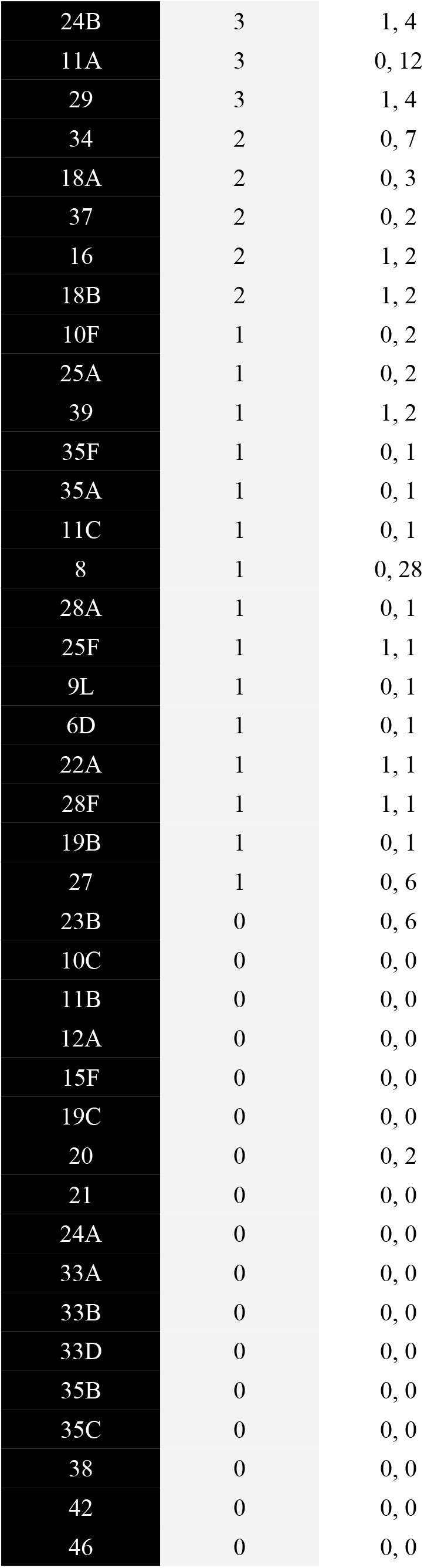
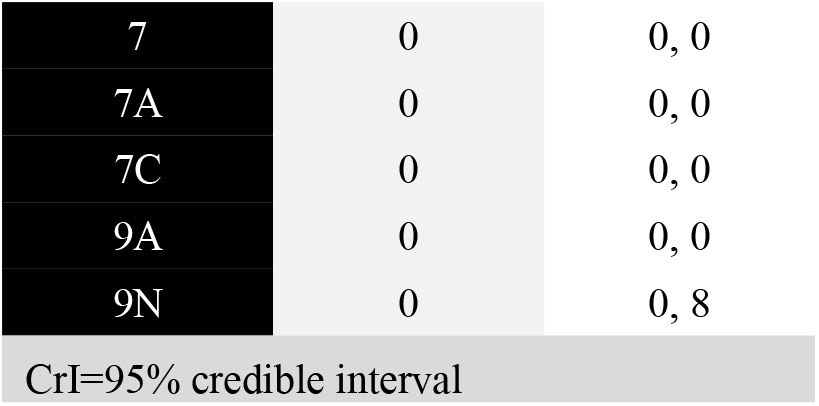
Estimated additional cases gained that are not explained by increased carriage in children. The median and 95% credible intervals for the unexplained excess cases during the full time period and across all ages is shown for each serotype.

| Serotype | Median | CrI |
| --- | --- | --- |
| 12F | 156 | 50, 268 |
| 24F | 53 | 25, 80 |
| 22F | 42 | 16, 69 |
| 10A | 30 | 19, 47 |
| 15BC | 22 | 5, 36 |
| 6C | 19 | 16, 22 |
| 16F | 19 | 0, 55 |
| 15A | 19 | 2, 39 |
| 33F | 18 | 0, 53 |
| 2 | 14 | 0, 23 |
| 10B | 13 | 3, 25 |
| 31 | 6 | 1, 20 |
| 13 | 6 | 1, 10 |
| 7B | 5 | 1, 16 |
| 23A | 5 | 0, 13 |
| 17F | 3 | 0, 10 |
| 24B | 3 | 1, 4 |
| 11A | 3 | 0, 12 |
| 29 | 3 | 1, 4 |
| 34 | 2 | 0, 7 |
| 18A | 2 | 0, 3 |
| 37 | 2 | 0, 2 |
| 16 | 2 | 1, 2 |
| 18B | 2 | 1, 2 |
| 10F | 1 | 0, 2 |
| 25A | 1 | 0, 2 |
| 39 | 1 | 1, 2 |
| 35F | 1 | 0, 1 |
| 35A | 1 | 0, 1 |
| 11C | 1 | 0, 1 |
| 8 | 1 | 0, 28 |
| 28A | 1 | 0, 1 |
| 25F | 1 | 1, 1 |
| 9L | 1 | 0, 1 |
| 6D | 1 | 0, 1 |
| 22A | 1 | 1, 1 |
| 28F | 1 | 1, 1 |
| 19B | 1 | 0, 1 |
| 27 | 1 | 0, 6 |
| 23B | 0 | 0, 6 |
| 10C | 0 | 0, 0 |
| 11B | 0 | 0, 0 |
| 12A | 0 | 0, 0 |
| 15F | 0 | 0, 0 |
| 19C | 0 | 0, 0 |
| 20 | 0 | 0, 2 |
| 21 | 0 | 0, 0 |
| 24A | 0 | 0, 0 |
| 33A | 0 | 0, 0 |
| 33B | 0 | 0, 0 |
| 33D | 0 | 0, 0 |
| 35B | 0 | 0, 0 |
| 35C | 0 | 0, 0 |
| 38 | 0 | 0, 0 |
| 42 | 0 | 0, 0 |
| 46 | 0 | 0, 0 |
| 7 | 0 | 0, 0 |
| 7A | 0 | 0, 0 |
| 7C | 0 | 0, 0 |
| 9A | 0 | 0, 0 |
| 9N | 0 | 0, 8 |
| CrI=95% credible interval |  |  |

## DISCUSSION

With several higher-valency conjugate vaccines under development, there is a need to understand and predict the patterns of serotype replacement to anticipate future changes. In this study, we created a model that quantifies changes in serotype prevalence post-PCV introduction in Israel based on carriage data. We found that the common assumption that non-vaccine serotypes increase by the same proportion underestimates changes in serotype prevalence in Jewish children and overestimates them in Bedouin children. The change in prevalence post-vaccination was too variable across serotypes to use only one value to represent the proportional change for all serotypes. Instead, we found that the change in the prevalence of serotypes over time is not the same for all serotypes but has a positively skewed distribution. Using pre-vaccine prevalence data, we can estimate where on the distribution of post-vaccine changes that serotypes will fall. We can then combine observed IPD estimates and model-estimated prevalence ratios to get an estimate of serotype-specific replacement due to increased carriage. Using the estimated prevalence ratios, along with observed IPD data, we were able to rank serotypes based on cases gained as a result of increased carriage in children. However, there were also additional cases gained over this study period that were not predicted by increased carriage, suggesting the need to include additional mechanisms in the estimates.

The estimated number of cases that were not explained by the increased carriage in children illuminated some of the limitations of using carriage alone to predict disease. Changes in carriage prevalence explained some of the variations in IPD in adults, but not all. A large number of excess IPD cases occurred during this time period that were not predicted by increased carriage. Serotype 12F in particular had a large number of unexplained cases gained. Surprisingly, serotype 8 did not have many unexplained cases. In fact, we would have expected there to be more IPD cases due to serotype 8 overall, either explained or unexplained.

Changes like serotype-specific epidemics cycles or changes in ascertainment could account for some of this variability. Moreover, carriage patterns in specific age groups (e.g., older children, adults) might better reflect the changes in IPD seen in these other age groups. We had intended to carry out this comparison, but our data were too sparse in subsets of the population. Future work could investigate this difference further.

We found that pre-vaccine prevalence was correlated with where on the distribution of slopes serotypes are located, with serotypes having higher pre-vaccine prevalence associated with larger increases after vaccination. In other words, more prevalent serotypes before introduction of PCV-13 were associated with larger increases over the study period.

Our method of ranking serotypes varied between age groups, but overall the same serotypes had the highest number of cases gained through serotype replacement after vaccination. Unsurprisingly, serotype 8 was among the highest serotypes in this ranking system. It has recently been seen to be one of the most important serotypes to emerge in other populations, particularly in older age groups, which is consistent with our findings. Also not surprising were the high rankings of serotypes 12F and 33F, which have been noted to increase in recent years.

Recent work has shown that models that incorporate genomics or clonal groups can forecast which serotypes will be successful after vaccination [19, 20, 31]. Future work could combine the results of this study with new genomics studies, directly incorporating information on the fitness of a serotype (e.g., as calculated by Azarian) in these models, or this type of model structure could be used to benchmark alternative modeling approaches.

A fundamental limitation of using carriage data to forecast changes in IPD is that changes in serotypes that are rarely carried in children cannot be easily tracked using carriage data. It is not clear if serotypes with these characteristics, like serotypes 1, 5, and 8 might be transmitted directly among older age groups (bypassing children) or if they might be transmitted between symptomatic individuals.

This study had some other limitations. Many serotypes are rare, and as a result we did not have estimates for all 90+ serotypes for all variables. For the carriage data, we had count data for only 67 serotypes. However, this lack of data for the other serotypes is not likely to impact the results greatly because they are so rare in both carriage and disease and would probably not factor into the model.

This analysis was limited to the available data in a single country. The dataset contained only seven years of data, and only included carriage data from children. The model structure additionally had some assumptions. We attempted to address these structural assumptions by comparing several models (variations in random and fixed effects, covariance matrices, predictors, and time). When compared to the observed data, the model appeared to fit, suggesting that these assumptions were valid. Future research could investigate these assumptions further, using other populations and additional time periods. Additionally, the IPD data available were comprised of mostly Jewish individuals. As a result, we were unable to extrapolate further for the Bedouin population past fitting the carriage model. Further analyses could explore higher transmission populations such as the Bedouin population.

Though we identified some of the top serotypes using our method of ranking, it is important to note that looking at individual serotypes can be sometimes misleading because the data are so noisy. Previous work demonstrates that estimating changes for specific serotypes can be subject to a high degree of noise, sometimes leading to inaccurate projections [17]. Future work could incorporate additional predictors of serotype prevalence growth as they become available.

Having a strong understanding of the patterns of serotype replacement could be important as newer higher-valent vaccines are developed and introduced. With higher-valency vaccines, we need to be able to predict both the decline in vaccine-serotypes and the increase in non-vaccine-serotypes. We can use this model to quantify changes in serotype replacement in other populations using the same analysis. This model may also help to optimize future serotype compositions.

## Data Availability

All code and simulated versions of the data are available online at https://github.com/mailephillips/post-pcv-expansion. Real data were not available for public use, so data were simulated for the purposes of sharing the code online.

## SUPPLEMENTAL INFORMATION

### S1 Text. Carriage Model

**Figure S1. Observed and expected prevalence plot, by year (Jewish children).** *Each panel shows the observed (black points) and model-fitted estimates of prevalence (different-colored dots and bars represent point estimates and 95% credible intervals), by year of the study period*.

**Figure S2. Observed and expected prevalence plot, by year (Bedouin children).** *Each panel shows the observed (black points) and model-fitted estimates of prevalence (different-colored dots and bars represent point estimates and 95% credible intervals), by year of the study period*.

**Figure S3. Observed versus fitted prevalence ratios (Jewish children).** *The plot shows the observed (black points) and model-fitted prevalence ratios (different-colored dots and bars represent point estimates and 95% credible intervals). The size of the different-colored dots indicates the number of isolates over the study period from the raw data (smaller dots = less isolates, larger dots = more isolates). Ratios shown are for the first to last years of the study period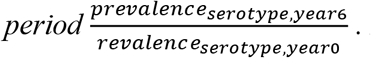. In instances where the observed prevalence in both Year 0 and Year 6 were 0, the observed prevalence ratio is indicated to be one. In cases where the observed prevalence in Year 0 was 0 and but the observed prevalence in Year 6 was nonzero, no observed point is shown (serotypes 20, 27, 29, and 7B)*.

**Figure S4. Observed versus fitted prevalence ratios (Bedouin children).** *The plot shows the observed (black points) and model-fitted prevalence ratios (different-colored dots and bars represent point estimates and 95% credible intervals). The size of the different-colored dots indicates the number of isolates over the study period from the raw data (smaller dots = less isolates, larger dots = more isolates). Ratios shown are for the first to last years of the study period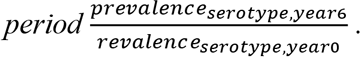*.

**Figure S5. Estimates for log relative risk ratios for pre-PCV period.** *Each serotype-specific log relative risk ratio with its 95% credible interval is shown for the pre-PCV period (*β_0,i_*), estimated from the model. The serotypes are shown from highest to lowest log relative risk ratios, separately for Jewish (A) and Bedouin (B) children*.

**Figure S6. Estimates for log relative risk ratios for early post-PCV period.** *Each serotype-specific log relative risk ratio with its 95% credible interval is shown for the early post-PCV period (*β_1,i_*), estimated from the model. The serotypes are shown from highest to lowest log relative risk ratios, separately for Jewish (A) and Bedouin (B) children*.

**Figure S7. Estimates for log relative risk ratios for late post-PCV period.** *Each serotype-specific log relative risk ratio with its 95% credible interval is shown for the late post-PCV period (*β_1,i_*), estimated from the model. The serotypes are shown from highest to lowest log relative risk ratios, separately for Jewish (A) and Bedouin (B) children*.

**Figure S8. Estimated additional cases gained as a result of increased carriage in children, by serotype.** *Each panel shows an individual serotype over the study period for all ages. Each full bar represents the total number of observed IPD cases for that stratum. The different colors denote the fraction of total cases attributable to different groups of serotypes: expected non-vaccine-targeted serotype cases had PCV13 not been introduced (solid red), estimated additional cases gained that are not explained by this model (hatched blue), and the estimated additional cases gained as a result of increased carriage in children (hatched red)*.

**Table S1. Penalized deviance for all model combinations fit.** *The deviance information criterion (DIC) used to determine the best fit model is shown for all combinations of models for both Jewish and Bedouin datasets. An “X” indicates whether that particular feature was used in building the specified model*.

**Table S2. Serotype-specific parameters estimated from model: Jewish children under 5 in Israel, November 2009-July 2016.** *Estimates for the serotype-specific pre-PCV relative risk ratios (RRRs) (*e^β_0,i_^*), early post-PCV RRRs (*e^β_1,i_^*), and late post-PCV RRRs (*e^β_2,i_^*) are shown for the Jewish carriage model, with their 95% credible intervals*.

**Table S3. Serotype-specific parameters estimated from model: Bedouin children under 5 in Israel, November 2009-July 2016.** *Estimates for the serotype-specific pre-PCV relative risk ratios (RRRs) (*e^β_0,i_^*), early post-PCV RRRs (*e^β_1,i_^*), and late post-PCV RRRs (*e^β_2,i_^*) are shown for the Bedouin carriage model, with their 95% credible intervals*.

